# Malaria parasitemia after mass distribution of azithromycin to prevent child mortality in Burkina Faso: results from a cluster randomized trial

**DOI:** 10.1101/2025.04.26.25326479

**Authors:** Boubacar Coulibaly, Ali Sié, Mamadou Ouattara, Mamadou Bountogo, Guillaume Compaoré, Adama Compaoré, Moustapha Nikiema, Nestor Dembélé Sibiri, Jérôme Nankoné Tiansi, Elodie Lebas, Ian Fetterman, Huiyu Hu, Thuy Doan, Benjamin F. Arnold, Thomas M. Lietman, Catherine E. Oldenburg

## Abstract

Twice-yearly mass distribution of azithromycin to children aged 1-59 months reduces all-cause child mortality. Some studies have suggested that mass azithromycin distributions may reduce malaria mortality and parasitemia, however these studies have been done in the absence of seasonal malaria chemoprevention (SMC). Here, we evaluated malaria parasitemia in a cluster randomized trial of azithromycin versus placebo in Burkina Faso that was receiving SMC. Thin and thick smears were taken from a random sample of 15 children per cluster in 40 clusters that had been receiving twice-yearly azithromycin or placebo for 36 months (6 distributions). We found no evidence of a difference in malaria parasitemia in children in azithromycin compared to placebo clusters (mean difference -6% prevalence, 95% CI -17% to 6%, *P*=0.33). These results suggest that reductions in malaria parasitemia may not be a major contributor to the effect of azithromycin on child mortality in settings receiving SMC.

## INTRODUCTION

Biannual mass distribution of azithromycin to children aged 1 to 59 months reduces all-cause child mortality in the Sahel.^1–3^ Previous studies in Niger have suggested that mass distribution of azithromycin may lead to decrease is malaria mortality and malaria parasitemia, suggesting that a reduction in malaria transmission may contribute to observed effects on all-cause mortality.^4,5^ Azithromycin has some activity against the plasmodial apicoplast^6,7^, and although it has not been found to reduce malaria parasitemia in individually-randomized trials^8,9^, community-wide distribution may reduce malaria transmission that could lead to population-level declines in prevalence. Here, we evaluate malaria parasitemia in children aged 1-59 months living in communities that received twice-yearly mass distribution of azithromycin compared to placebo for 36 months (6 distributions). We hypothesized that children living in communities receiving azithromycin would have lower prevalence of malaria parasitemia compared to placebo.

## METHODS

This study was conducted in a subset of communities participating in the Child Health with Azithromycin Treatment (CHAT) trial, a 1:1 randomized placebo-controlled trial of twice-yearly azithromycin distribution to children aged 1-59 months compared to placebo for prevention of child mortality.^1,10^ The primary outcome for the trial was all-cause mortality and has been previously reported.^1^ Complete methods have previously been reported.^1,10^ CHAT was conducted in Nouna District, Burkina Faso, located in northwestern Burkina Faso, from 2019 to 2023. Nouna received four monthly rounds of seasonal malaria chemoprevention (SMC) per year from July through October during the trial. Nouna District includes the Nouna Health and Demographic Surveillance Site (HDSS)^11^; all communities in the district were eligible for inclusion in the trial. Communities with more than 2,000 residents were split into multiple randomization units. Communities were randomized 1:1 to mass distribution of a single, oral dose of azithromycin or matching placebo to all children aged 1-59 months. Participants, investigators, and study staff were masked to treatment allocation. At baseline prior the start of trial activities, a random sample of 48 randomization units from within the HDSS were selected to participate in morbidity monitoring, including collection of thin and thick smears for malaria measurement. Samples were collected at baseline prior to any treatment distribution and at 36 months, after communities had received 6 rounds of azithromycin or placebo. Fifteen children per community were randomly selected from the most recent study census for sample collection. Assuming parasitemia prevalence of 20% in the placebo arm and an intracluster correlation of 0.056, this sample size was estimated to provide at least 80% power to detect a difference of 8.3% between the arms.

Baseline exams were conducted from August through November 2019. At 36 months (December 2022 through February 2023), some of the original clusters were not accessible due to security concerns. We randomly selected replacement clusters that were participating in the trial but not in the original morbidity monitoring. The number of replacement clusters was based on the number of accessible clusters at the time of the study.

Thin and thick smears were collected for assessment of malaria parasitemia at baseline and 36 months from each child selected for examination in each community. Blood smears were collected on glass slides, air dried, and stored at room temperature. Slides reading was done by qualified microscopists and according to the malaria microscopy protocol at the Centre de Recherche en Santé de Nouna. In brief slides were stained with 3% Giemsa for 30 minutes and read by two microscopists who were masked to each other’s grade. The microscopists determined the presence or absence of *Plasmodium* spp parasites and number of sexual and asexual parasites per μl (assuming 8000 white blood cells per μl) to assess parasite density.

Only asexual parasites density was considered for analysis. A positive smear was considered positive for malaria parasitemia, and slides with discrepant results by the two microscopists were adjudicated by a third masked microscopist. The mean of the two parasite density measurements was used for analysis.

The pre-specified analysis plan for the malaria outcome specified a linear regression model conducted at the level of the randomization unit (the cluster) with cluster-level prevalence as the outcome with the cluster’s randomized treatment assignment as the sole predictor. Parasite density was assessed using a linear regression model at the cluster level, with the mean cluster-level parasite density as the outcome and the cluster’s randomized treatment assignment as the sole predictor. A second non-prespecified secondary analysis included adjustment for baseline measures in the 25 communities that had both baseline and 36-month parasitemia measurements using a similar analysis strategy as the primary outcome with an additional covariate for baseline parasitemia. All analyses were run in R (The R Foundation for Statistical Computing, Vienna, Austria).

## RESULTS

At baseline, samples were collected from 613 children in 41 communities, including 23 in the azithromycin group (N=345 children) and 18 in the placebo group (N=268 children; **Figure 1**). At 36 months, samples were collected from 689 children in 40 communities (20 communities and 345 children in the azithromycin group and 20 communities and 344 children in the placebo group; **Figure 1**). Approximately 48% of children were female and median age was 23 months (interquartile range 14 to 36). At baseline, the prevalence of malaria was 14% in the azithromycin group and 21% in the placebo group (**Table 1**).

**Table 1.** Baseline characteristics.

|  | **Azithromycin** | **Placebo** |
| --- | --- | --- |
| N communities | 23 | 18 |
| N children per community, mean (95% CI) | 143 (109, 176) | 144 (109, 179) |
| N children sampled per community, mean (95% CI) | 15 (15, 15) | 15 (15, 15) |
| % female, mean (95% CI) | 48% (44, 52) | 48% (41, 53) |
| Age, % |  |  |
| 0 y | 20.87% | 17.91% |
| 1 y | 35.94% | 25.00% |
| 2 y | 19.71% | 26.12% |
| 3 y | 12.75% | 14.93% |
| 4 y | 10.72% | 16.04% |
| Baseline parasitemia, mean (95% CI) | 14.2% (9.7 to 18.7%) | 20.8% (12.7 to 28.9%) |

**Figure 1.**
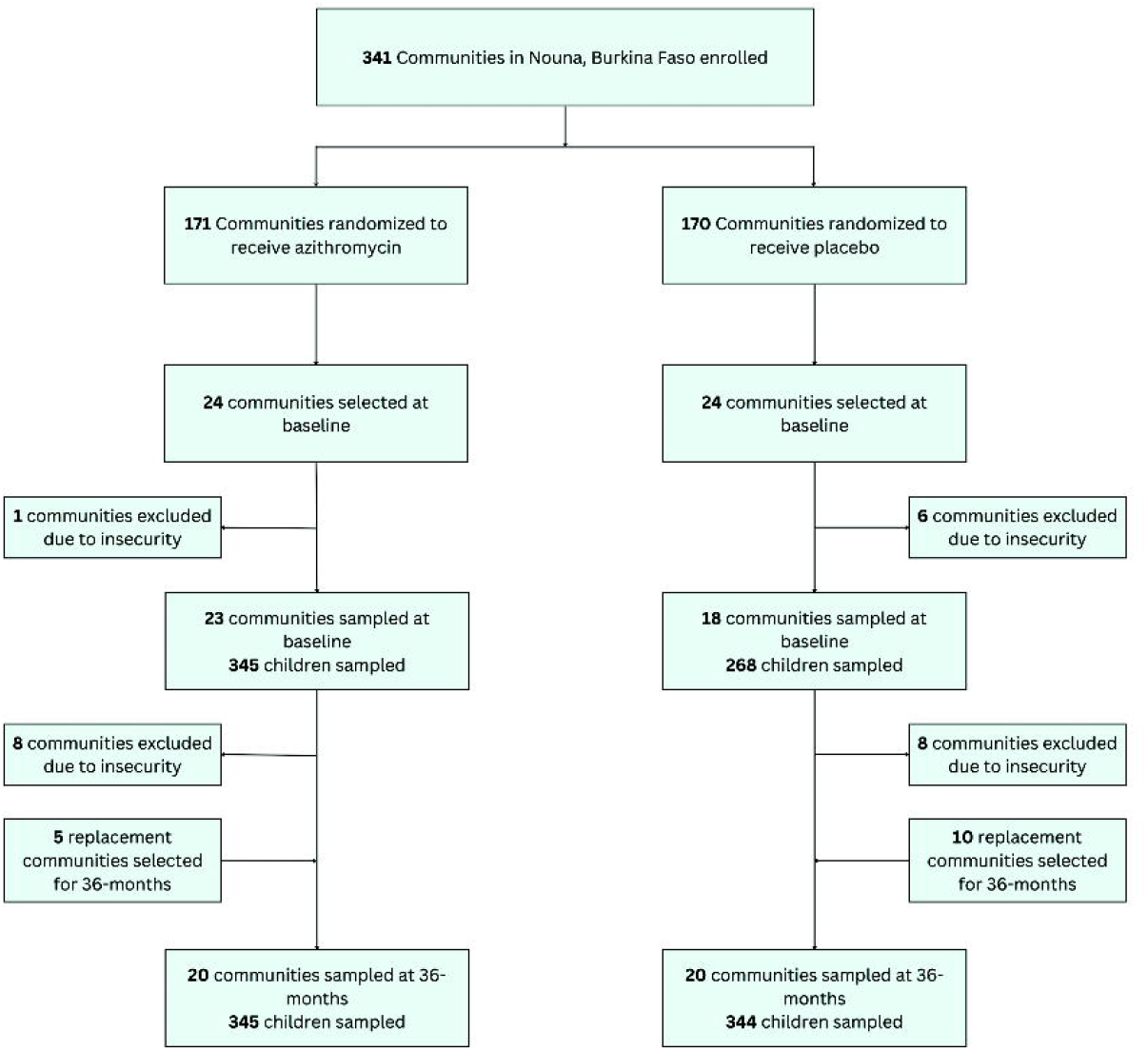
CONSORT diagram for communities included in malaria outcome monitoring.

At 36 months, malaria parasitemia prevalence was 26% in the azithromycin group (95% confidence interval, CI, 19 to 33%) and 32% in the placebo group (95% CI 22 to 41%), corresponding to a mean difference of 6% (95% CI -17 to 6%, *P*=0.33; **Table 2**). Mean square root parasite density was 40.8 parasites/μl in the azithromycin group (95% CI 18.1 to 63.6 parasites/μl) and 54.3 parasites/μl in the placebo group (95% CI 21.5 to 87.2 parasites/μl, mean difference 13 parasites/μl, *P*=0.48).

**Table 2.**
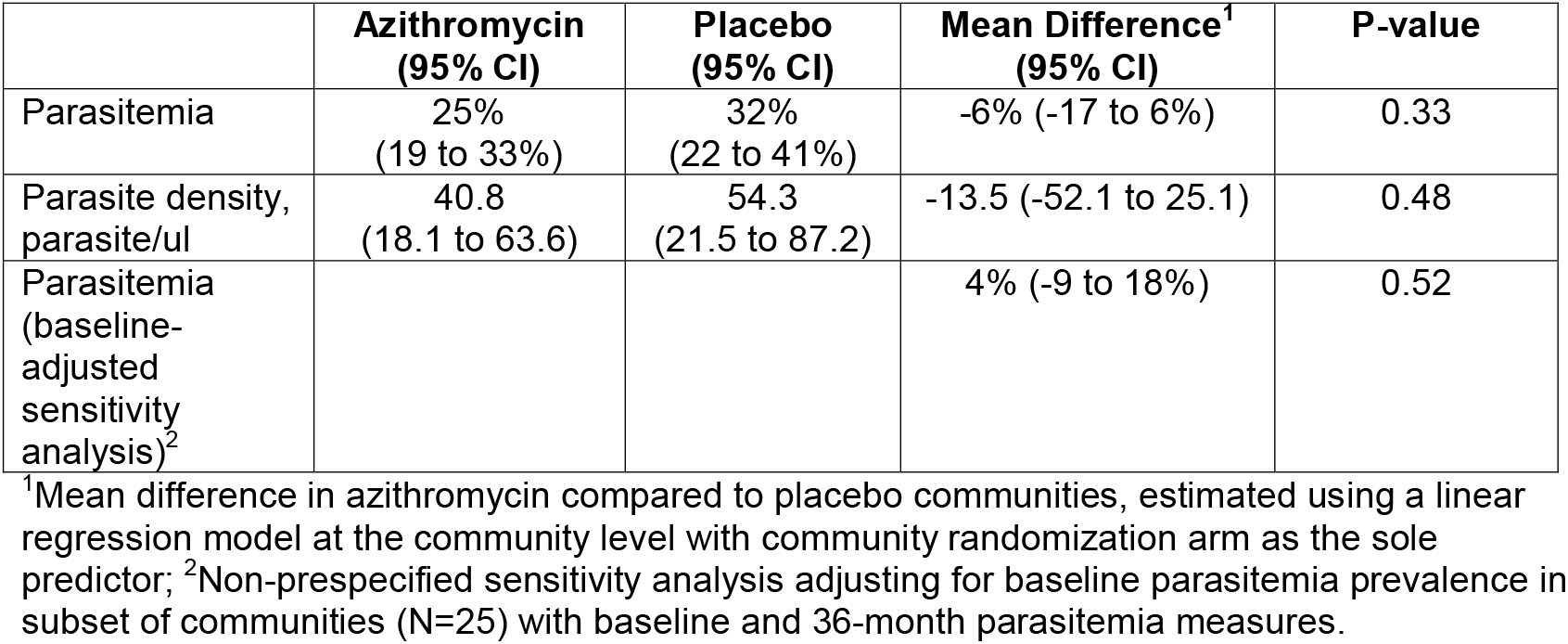
Community prevalence of malaria parasitemia at 36 months.

|  | **Azithromycin**<br>**(95% CI)** | **Placebo**<br>**(95% CI)** | **Mean Difference<sup>1</sup>**<br>**(95% CI)** | **P-value** |
| --- | --- | --- | --- | --- |
| Parasitemia | 25%<br>(19 to 33%) | 32%<br>(22 to 41%) | -6% (-17 to 6%) | 0.33 |
| Parasite density, parasite/ul | 40.8<br>(18.1 to 63.6) | 54.3<br>(21.5 to 87.2) | -13.5 (-52.1 to 25.1) | 0.48 |
| Parasitemia (baseline-adjusted sensitivity analysis)<sup>2</sup> |  |  | 4% (-9 to 18%) | 0.52 |

<sup>1</sup>Mean difference in azithromycin compared to placebo communities, estimated using a linear regression model at the community level with community randomization arm as the sole predictor; <sup>2</sup>Non-prespecified sensitivity analysis adjusting for baseline parasitemia prevalence in subset of communities (N=25) with baseline and 36-month parasitemia measures.

In a non-prespecified analysis of 25 clusters that had baseline and 36-month data, we found no evidence of a difference in malaria parasitemia in children in communities receiving azithromycin compared to placebo adjusting for baseline malaria parasitemia (mean difference 4%, 95% CI -9 to 18%, *P*=0.51; **Table 2**).

## DISCUSSION

We found no evidence of a difference in malaria parasitemia in children aged 1-59 months in communities in Burkina Faso receiving twice-yearly azithromycin distribution compared to placebo. In Niger, studies have found modest reductions in malaria parasitemia and malaria mortality among children living in communities receiving azithromycin compared to placebo.^4,5^ One important difference between the current study and the previous study in Niger is that all communities in the current trial in Burkina Faso received SMC during the high malaria transmission season. A previous household randomized trial of SMC with or without azithromycin found no evidence of a difference in malaria parasitemia in children receiving SMC plus azithromycin compared to SMC plus placebo.^12^ In the presence of SMC, the mild antimalarial activity of azithromycin may not be sufficient to lead to population-level decreases in parasitemia. However, at 36 months, data were collected after the final SMC distribution in all study clusters, so this may not fully explain the lack of effect of mass azithromycin distribution on malaria parasitemia in this setting.

This study has several limitations. Due to an evolving security situation in Burkina Faso, not all communities were accessible for sample collection at baseline and 36 months, reducing the overall sample size and resulting in lack of baseline data in some communities. Baseline malaria parasitemia was higher in the placebo group compared to the azithromycin group. A sensitivity analysis adjusting for baseline in a subset of communities with measures at both timepoints did not affect conclusions. The study was powered for approximately an 8% difference in parasitemia prevalence, and results were consistent with a 6% reduction, with relatively wide confidence intervals. The study was likely underpowered to detect potentially small but meaningful differences in parasitemia prevalence. Data were collected at endline (36 months) only, approximately 6 months after the most recent treatment distribution; this study was unable to demonstrate any shorter-term effects of azithromycin on malaria parasitemia.

Malaria transmission is highly seasonal in Burkina Faso, with transmission declining in January following cessation of the rainy season. Data were collected over a 4-month period at 36 months, and thus malaria parasitemia prevalence likely declined over time. The sample size was not sufficient to assess any differential effect of azithromycin over time, however sample collection was similar by arm by month, and the trial was fully masked, so data collectors and microscopists were not aware of the clusters’ treatment assignments. All samples at 36 months were collected after the SMC season, and we cannot comment on whether malaria parasitemia prevalence would be different in azithromycin versus placebo communities at the time of SMC distribution.

We found no evidence of a difference in malaria parasitemia prevalence among young children living in communities receiving twice yearly mass azithromycin distribution compared to placebo for prevention of child mortality. Although CHAT found an 18% reduction in child mortality in communities receiving azithromycin compared to placebo^1^, these results suggest that reductions in malaria may not be the primary mechanism of this reduction.

## Data Availability

All data produced in the present study are available upon reasonable request to the authors.

## FUNDING

The CHAT trial was supported by the Bill and Melinda Gates Foundation (OPP1187628, PI: Lietman). The funder played no role in the study design, data collection, interpretation of data, or decision to publish.

## AUTHOR CONTACT INFORMATION

Boubacar Coulibaly

Ali Sié

Mamadou Ouattara

Mamadou Bountogo

Guillaume Compaoré

Adama Compaoré

Moustapha Nikiema

Nestor Dembélé Sibiri

Jérôme Nankoné Tiansi

Elodie Lebas

Ian Fetterman

Huiyu Hu

Thuy Doan

Benjamin F. Arnold

Thomas M. Lietman

Catherine E. Oldenburg

## Notes

### Competing Interest Statement

The authors have declared no competing interest.

### Clinical Trial

NCT03676764

### Author Declarations

The institutional review board of the University of California, San Francisco and the Comite d'Ethique pour la Recherche en Sante in Ouagadougou, Burkina Faso gave ethical approval for this work.

## REFERENCES

1 Oldenburg C, Ouattara M, Bountogo M, et al. Mass Azithromycin Distribution to Prevent Child Mortality in Burkina Faso: The CHAT Randomized Clinical Trial. JAMA2024; 331: 482–90.

2 Keenan JD, Bailey RL, West SK, et al. Azithromycin to Reduce Childhood Mortality in Sub-Saharan Africa. New England Journal of Medicine2018; 378: 1583–92.

3 O’Brien KS, Arzika AM, Amza A, et al. Azithromycin to Reduce Mortality — An Adaptive Cluster-Randomized Trial. New England Journal of Medicine2024; 391: 699–709.

4 Keenan JD, Arzika AM, Maliki R, et al. Cause-specific mortality of children younger than 5 years in communities receiving biannual mass azithromycin treatment in Niger: verbal autopsy results from a cluster-randomised controlled trial. Lancet Global Health2020; 8: 288–95.

5 Arzika AM, Maliki R, Boubacar N, et al. Biannual mass azithromycin distributions and malaria parasitemia in pre-school children in Niger: A cluster-randomized, placebo-controlled trial. PLoS Med2019; 16: e1002835.

6 Sidhu ABS, Sun Q, Nkrumah LJ, Dunne MW, Sacchettini JC, Fidock DA. In Vitro Efficacy, Resistance Selection, and Structural Modeling Studies Implicate the Malarial Parasite Apicoplast as the Target of Azithromycin. J Biol Chem2007; 282: 2494–504.

7 Dahl EL, Rosenthal PJ. Multiple Antibiotics Exert Delayed Effects against the Plasmodium falciparum Apicoplast. Antimicrob Agents Chemother2007; 51: 3485–90.

8 Coulibaly B, Sié A, Dah C, et al. Effect of a single dose of oral azithromycin on malaria parasitaemia in children: a randomized controlled trial. Malar J2021; 20: 1–8.

9 Brogdon J, Dah C, Sié A, et al. Malaria positivity following a single oral dose of azithromycin among children in Burkina Faso: a randomized controlled trial. BMC Infect Dis2022; 22. DOI:10.1186/s12879-022-07296-4.

10 Sié A, Ouattara M, Bountogo M, et al. A double-masked placebo-controlled trial of azithromycin to prevent child mortality in Burkina Faso, West Africa: Community Health with Azithromycin Trial (CHAT) study protocol. Trials2019; 20: 675.

11 Sie A, Louis VR, Gbangou A, et al. The Health and Demographic Surveillance System (HDSS) in Nouna, Burkina Faso, 1993–2007. Glob Health Action2010; 3: 5284.

12 Chandramohan D, Dicko A, Zongo I, et al. Effect of Adding Azithromycin to Seasonal Malaria Chemoprevention. New England Journal of Medicine2019; 380: 2197–206.

